# A spectrum of *BRCA1* and *BRCA2* germline deleterious variants in ovarian cancer in Russia

**DOI:** 10.1101/2021.07.22.21260831

**Authors:** Andrey Kechin, Ulyana Boyarskikh, Alexey Barinov, Alexander Tanas, Svetlana Kazakova, Anastasia Zhevlova, Evgeniy Khrapov, Sergey Subbotin, Olga Mishukova, Tatiana Kekeeva, Irina Demidova, Maxim Filipenko

## Abstract

Pathogenic mutations in *BRCA1* and *BRCA2* genes are essential biomarkers of an increased breast and ovarian cancer risk and tumor sensitivity to poly ADP ribose (PARP) inhibitors. In many countries, their detection includes patient prescreening with quantitative PCR (qPCR) identifying several founder mutations. In Russia, eight mutations are included in such tests, among which c.5266dup (5382insC) is the most frequently identified one. Here, we showed the distribution of 1406 pathogenic or likely pathogenic mutations in *BRCA1/2* genes identified in ovarian cancer patients recruited into the study from 72 Russian regions in 2015-2021. The most of mutations were detected with qPCR, for qPCR mutation negative samples, targeted next-generation sequencing (NGS) covering whole coding sequences of the genes was applied. As expected, the most abundant mutations were c.5266dupC (41.0%), c.4035delA (7.0%), c.1961delA (6.3%), c.181T>G (5.2%), c.3756_3759delGTCT (1.8%), c.3700_3704delGTAAA (1.5%), and c.68_69delAG (1.5%). However, we identified several mutations which were more frequent than the founder c.5946delT mutation (also known as 6174delT, 0.5% of participants): c.5152+1G>T (1.2%), c.1687C>T (1.0%), c.4689C>G (0.9%), c.1510delC (0.6%), c.2285_2286delGA (0.6%) in the *BRCA1* gene; and c.5286T>G (1.2%), c.2808_2811delACAA (0.8%), c.658_659delGT (0.7%), c.7879A>T (0.6%), c.3847_3848delGT (0.6%) in the *BRCA2* gene. Having developed an NGS-targeted panel on SNPs flanking *BRCA2* c.5286T>G, we showed the founder effect for this mutation and suggested that it arose about 700 years ago that is twice later that it is thought for the c.5266dupC. The total occurrence of mutations identified in at least 10 participants (13 mutations) was only 70%. To our knowledge, eighty-nine mutations (identified in 8% of participants) have not been described previously. Thus, this study may help in improving prescreening qPCR tests and extend our knowledge about the *BRCA1* and *BRCA2* genes variability in ovarian cancer patients.

## Introduction

*BRCA1* and *BRCA2* are the genes mutated frequently in ovarian cancer patients [1]. Their influence on genome maintenance is associated with their essential role in the repair of double-strand breaks. This link has led to the development of new targeted therapy with poly ADP ribose polymerase (PARP) inhibitors that show more effective and less toxic treatment than common chemotherapy [2]. Several such drugs have been approved for use by the U.S. Food and Drug Administration (FDA) (olaparib, rucaparib, and niraparib), and some new are in the late stage of clinical development [3]. The main indication for the use of such drugs is the presence of pathogenic variants in the *BRCA1* or *BRCA2* gene, therefore their occurrence in a population is the necessary information for the effective organizing of the ovarian cancer patients’ testing. Today, more than 6500 pathogenic and likely pathogenic variants are known for the *BRCA1* and *BRCA2* genes according to the ClinVar database, and this number increases each year. The occurrence of the pathogenic and likely pathogenic variants varies between different populations significantly, particularly in their representation among all *BRCA1/2* pathogenic variant carriers. The ten most common variants can be from 33 to 89 % of all carriers [4–9] depending on the population studied. For populations with a high contribution of founder mutations, population prescreening with simple methods like qPCR can be applied as it has been suggested for the USA Ashkenazi patients over many years [10].

The *BRCA1/2* pathogenic variant frequency in Russia strongly deviates to several founder mutations (c.5266dupC, c.4153delA, and c.68_69delAG in the *BRCA1* and c.5946delT in the *BRCA2*). Recently studying the data obtained with PCR, Sanger, and next-generation sequencing showed that whole-coding *BRCA1/2* gene analysis in Russian patients can increase the number of mutation carriers identified twice [11]. However, the whole-coding data were obtained only for 785 patients with both breast and ovarian cancer, among which only 117 were mutation carriers. To evaluate the occurrence of founder mutations known and to discover new ones, we studied *BRCA1* and *BRCA2* genes with targeted NGS and quantitative PCR (qPCR) in 1406 ovarian cancer patients. To our knowledge, this is the largest *BRCA1/2* genes study for Russian populations.

## Materials and Methods

### Subjects

1406 ovarian cancer patients with revealed *BRCA1/2* pathogenic variant were recruited into the study by two main centers in Moscow and one in Novosibirsk in 2015-2021. The study was approved by the local medical ethics committee of the Institute of Chemical Biology and Fundamental Medicine of the Siberian Branch of the Russian Academy of Sciences. Among 1056 patients for which the region was known, patients were from 72 different regions. The most numbers were from Moscow (114), Primorsky Krai (88), Novosibirsk Oblast (76), and Moscow region (71). All participants provided informed consent for use of their genetic material in the study. DNA from blood leukocytes was extracted using an in-house method comprising cell lysis using 10% SDS-containing buffer, proteinase K treatment, protein extraction using phenol-chloroform, and isopropanol precipitation of the DNA.

### Studying BRCA1 and BRCA2 genes

The most of founder mutations (771 patients, *BRCA1*: c.5266dupC (5382insC), c.4035delA (4154delA), c.1961delA (2080delA), c.181T>G (C61G), c.3756_3759delGTCT (3875delGTCT), c.3700_3704delGTAAA (3819delGTAAA), c.68_69delAG (185delAG), *BRCA2*: c.5946delT (6174delT)) were identified with qPCR. For patients without hotspot mutation identified, the *BRCA1* and *BRCA2* coding sequences were studied using the in-house amplicon-based targeted NGS-panel (571 patients), GeneRead QIAact BRCA 1/2 panel (Qiagen) (64 patients). Coding exons, splice-acceptor, and splice-donor sites were covered (transcripts NM_007294.3 and NM_000059.3). NGS libraries were sequenced with MiSeq and MiniSeq Illumina platforms (351 patients), or with Ion S5/Ion Chef System (ThermoFisher) (284 patients). The NGS data was analyzed with the BRCA-analyzer [12] or Torrent Suite software followed by ANNOVAR annotation [13]. Visual data analysis, manual filtering of sequencing artifacts, and sequence alignments were performed using the Integrative Genomics Viewer (IGV) [14].

### Mutation age estimates

To estimate the age of some mutations for which we suggested the founder effect, we designed a new amplicon-based NGS-panel targeting SNPs flanking ± 5Mb around the *BRCA2* gene (28 SNPs with CEU population frequency ≥ 30%) with the NGS-PrimerPlex program [15] (Table 1). SNPs were chosen based on the minor allele frequency and the allele linkage disequilibrium (LD) from the LDProxy tool (https://ldlink.nci.nih.gov/). The next SNP was chosen in such a way that the LD with the previous one would be less than 0.7. Primer sequences are in Supplementary Table 1. Phased genotypes were obtained using the 1000Genomes [16] data and the Beagle tool [17]. Mutation ages were estimated with the Mutation dating online tool [18].

**Table 1.** SNPs that were covered by the targeted NGS panel to estimate mutation age. All positions are on chromosome 13 (based on the hg19 human genome assembly). The genome region studied is a chromosome part between the left and the right primers.

| dbSNP ID | Genome region studied start | Genome region studied end | Distance to the BRCA2 gene |
| --- | --- | --- | --- |
| rs201463755 | 27546887 | 27546913 | -5.36 Mb |
| rs9554211 | 28530006 | 28530030 | -4.38 Mb |
| rs661732 | 29528980 | 29529005 | -3.38 Mb |
| rs9508456 | 30016336 | 30016362 | -2.90 Mb |
| rs818608 | 30513048 | 30513069 | -2.40 Mb |
| rs35923108 | 31018930 | 31018960 | -1.89 Mb |
| rs4356336 | 31319525 | 31319561 | -1.60 Mb |
| rs1475283 | 31818878 | 31818898 | -1.09 Mb |
| rs12868737 | 32300812 | 32300843 | -0.61 Mb |
| rs798972 | 32737554 | 32737576 | -0.17 Mb |
| rs798989 | 32781283 | 32781318 | -0.13 Mb |
| rs12877839 | 32847808 | 32847838 | -0.06 Mb |
| rs206115 | 32888473 | 32888516 | -0.02 Mb |
| rs206319 | 32986209 | 32986243 | +0.03 Mb |
| rs1088550 | 33036646 | 33036671 | +0.08 Mb |
| rs2010997 | 33070479 | 33070506 | +0.11 Mb |
| rs9595795 | 33122795 | 33122817 | +0.17 Mb |
| rs2149859 | 33553601 | 33553650 | +0.60 Mb |
| rs7328041 | 33876950 | 33876971 | +0.92 Mb |
| rs9538629 | 34369034 | 34369052 | +1.42 Mb |
| rs7329029 | 34863088 | 34863114 | +1.91 Mb |
| rs1396310 | 35292999 | 35293014 | +2.34 Mb |
| rs2068462 | 35768034 | 35768045 | +2.81 Mb |
| rs9574293 | 36218034 | 36218062 | +3.26 Mb |
| rs9315379 | 36617462 | 36617482 | +3.66 Mb |
| rs751254 | 37088513 | 37088540 | +4.14 Mb |
| rs9576151 | 37570467 | 37570485 | +4.62 Mb |
| rs9532064 | 38053618 | 38053645 | +5.10 Mb |

## Results

### Deleterious mutations with high frequency

For 1406 patients positive for a deleterious *BRCA1/2* mutation, the occurrences of the most frequent (≥7 samples) mutations are presented in Table 2. The whole list of mutations identified is in Supplementary Table 2. For 1161 (82.6%) and 245 (17.4%) patients, the deleterious mutation was in the *BRCA1* or *BRCA2* gene, respectively, corresponding to 266 unique pathogenic variants, 128 (48%) and 138 (52%) in *BRCA1* and *BRCA2* genes, respectively. The number of variants in the ovarian cancer cluster region (OCCR) and the breast cancer cluster region (BCCR) [19] was 100 (37.2%) and 71 (26.4%) which corresponded to 426 (30.2%) and 762 (54.2%) participants, respectively.

**Table 2.** The most frequent germline deleterious mutations in the *BRCA1* and *BRCA2* genes found. ^*^ designates founder mutations in Europe [20]. The new mutation is in bold.

| Gene | CDS mutations | Type of mutation | Traditional name | Occ. | Ref. |
| --- | --- | --- | --- | --- | --- |
| *BRCA1* | c.5266dupC | frameshift variant | 5382insC | 577 * |  |
| *BRCA1* | c.4035delA | frameshift variant | 4154delA | 98 * |  |
| *BRCA1* | c.1961delA | frameshift variant | 2080delA | 89 * | [21] |
| *BRCA1* | c.181T>G | missense variant | C61G | 73 * |  |
| *BRCA1* | c.3756_3759delGTCT | frameshift variant | 3875delGTCT | 26 * |  |
| *BRCA1* | c.3700_3704delGTAAA | frameshift variant | 3819delGTAA<br>A | 21 * |  |
| *BRCA1* | c.68_69delAG | frameshift variant | 185delAG | 21 * |  |
| *BRCA2* | c.5286T>G | stop gained |  | 17 |  |
| *BRCA1* | c.5152+1G>T | splice donor variant |  | 17 |  |
| *BRCA1* | c.1687C>T | stop gained |  | 14 * |  |
| *BRCA1* | c.4689C>G | stop gained |  | 13 * | [22] |
| *BRCA2* | c.2808_2811delACAA | frameshift variant |  | 11 * | [23] |
| *BRCA1* | c.1510delC | frameshift variant |  | 9 |  |
| *BRCA2* | c.7879A>T | missense variant |  | 9 * | [24] |
| *BRCA2* | c.658_659delGT | frameshift variant |  | 9 * | [7,25] |
| *BRCA2* | c.3847_3848delGT | frameshift variant |  | 8 * |  |
| ***BRCA1*** | **c.2285_2286delGA** | frameshift variant |  | 8 |  |
| *BRCA2* | c.5946delT | frameshift variant | 6174delT | 7 * |  |

41.0% of all samples tested contained c.5266dupC, 7.0% – c.4035delA, 6.3% – c.1961delA, 5.2% – c.181T>G, 1.8% – c.3756_3759delGTCT, 1.5% – c.3700_3704delGTAAA, 1.5% – c.68_69delAG, 1.2% – c.5286T>G, 1.2% – c.5152+1G>T. c.5286T>G was previously considered as rare. Other mutations represented 33.2 % of all cases including 12.9 % corresponding to unique mutations. Totally, 89 new mutations (unknown in the ClinVar database on June 1, 2021) were identified in 113 participants, 36 in the *BRCA1* gene (51 participants), and 55 in the *BRCA2* gene (62 participants).

Due to the founder effect known for several mutations detected (c.5266dupC, c.4035delA, c.1961delA, c.181T>G), we evaluated the mutation occurrence ratios in different regions of Russia (Figure 1). For most regions with at least 20 pathogenic variant carriers, the c.5266dupC mutation occurred in 39–80 %, except for the Khanty-Mansi Autonomous Okrug (only 18% of 34 carriers) that could be associated with indigenous peoples living in the region. The total occurrence of the four founder mutations varied between 44 and 88 % depending on the region with the lowest values for the Khanty-Mansi Autonomous Okrug and the Chelyabinsk oblast.

**Figure 1.**
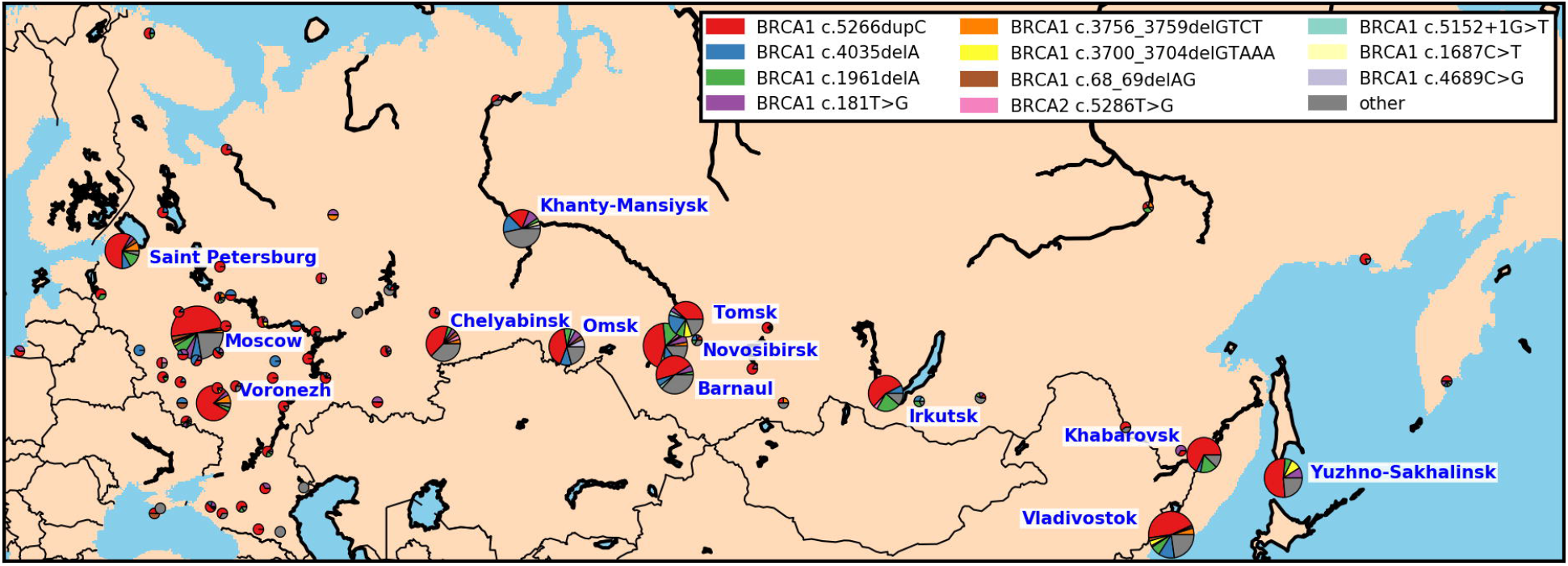
The mutation occurrence in different regions of Russia. The pie charts are shown in the regional administrative centers. For regions with more than 20 mutation carriers identified, the pie chart radiuses reflect the number of mutation carriers found. The threshold was chosen based on the convenience of the map element locations.

For some of the mutations detected, the founder effect was suggested earlier (Table 2), and others were found in several cases for the first time. Therefore, for all such mutations, we provided the literature review, and for the *BRCA2* c.5286T>G, we estimated the mutation age.

### New founder mutation BRCA2 c.5286T>G

*BRCA2* c.5286T>G was found in 17 ovarian cancer patients from different Russian regions (Moscow, Novosibirsk, Irkutsk, Kirov, and Chelyabinsk Oblasts, Yakutiya, Primorsky Krai) and the mutation common origin was suggested. To confirm it, we compared the *BRCA2* gene SNPs identified by sequencing *BRCA1/2* exons by a targeted NGS panel. For 6 patients, 12 flanking SNP phased alleles (from 32888483 to 32931875, hg19 human genome assembly) were the same. To estimate the mutation age, we developed a new small targeted NGS panel covering 28 SNPs around the *BRCA2* gene. For the moment of this study stage, the DNA samples were available only for six patients. The number of generations since the last common shared ancestor was determined as 34.8 (700 years, CI_95_: 19.6–62.2) and 41.1 (820 years, CI_95_: 19.6–62.2) for an independent and correlated genealogy, respectively. This mutation age is less than for the c.5266dupC determined earlier (about 72 generations) [26], and this value needs to be re-estimated for the higher number of samples and variations.

### Other BRCA2 highly recurrent mutations

Five *BRCA2* mutations found in this study have earlier been determined as founder ones in Europe: c.2808_2811delACAA (11 patients, found in Spain), c.658_659delGT (10 patients, Lithuania), c.7879A>T (9 patients, Macedonia), c.3847_3848delGT (8 patients, Denmark and Norway), c.5946delT (also known as 6174delT, 7 patients, Ashkenazi Jews and Hungary) (Table 3).

**Table 3.**
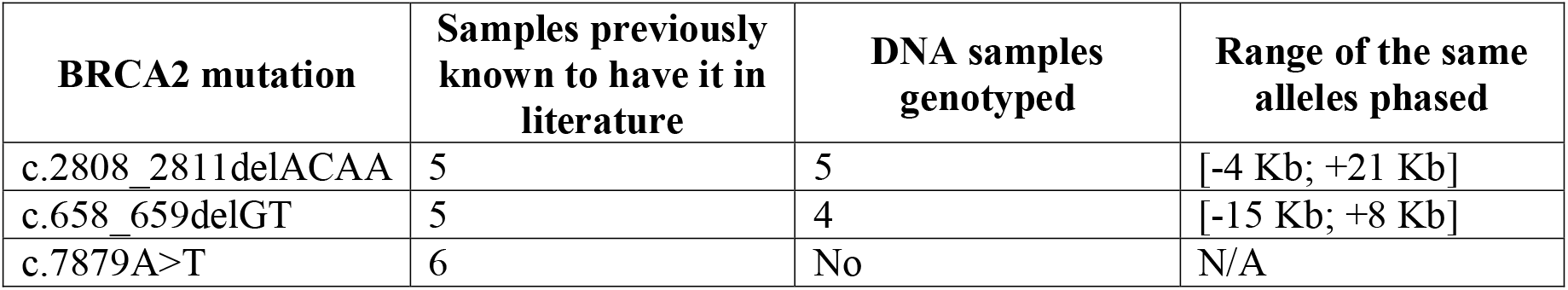
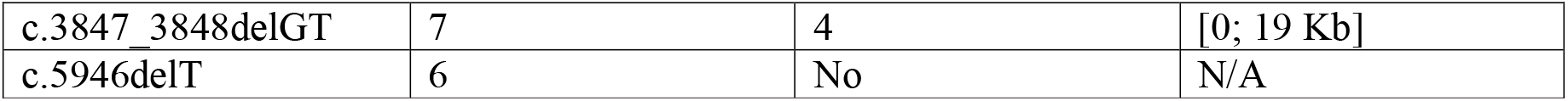
*BRCA2* mutations identified known to be a founder in other populations.

| **BRCA2 mutation** | **Samples previously known to have it in literature** | **DNA samples genotyped** | **Range of the same alleles phased** |
| --- | --- | --- | --- |
| c.2808_2811delACAA | 5 | 5 | [-4 Kb; +21 Kb] |
| c.658_659delGT | 5 | 4 | [-15 Kb; +8 Kb] |
| c.7879A>T | 6 | No | N/A |
| c.3847_3848delGT | 7 | 4 | [0; 19 Kb] |
| c.5946delT | 6 | No | N/A |

### BRCA1 highly recurrent mutations

Five *BRCA1* mutations were identified in at least eight patients: c.5152+1G>T (17 patients), c.1687C>T (14 patients), c.4689C>G (13 patients), c.1510delC (9 patients), and c.2285_2286delGA (8 patients). Due to the absence of the genotyping system, we couldn’t compare genotypes of the flanking SNPs for these *BRCA1* mutations. However, we reviewed thir occurrence in other studies. c.1687C>T is a known founder mutation in Austria, Slovenia, and Sweden [20]; c.4689C>G was earlier identified in many patients in Germany, USA, and Russia [22]. However, c.5152+1G>T was found only in two Russian patients [27,28] and in several families in the worldwide study [29]; c.1510delC was earlier found in some studies [30–32]; and c.2285_2286delGA is a new mutation. All of these mutations were observed more frequent than the *BRCA2* c.5946delT known to be founder and may be considered to be included into prescreening qPCR tests.

## Discussion

Here, we confirmed that *BRCA1* c.5266dupC was the most abundant germline pathogenic variant in Russian ovarian cancer patients accounting for up to 50 % of all *BRCA1/2* mutation cases. However, its frequency is significantly lower than 90%, as it was thought before the start of NGS application when most pathogenic variants were identified with qPCR [33]. Its high occurrence in Russia can be explained by the spreading from Scandinavia or northern Russia about 1800 years ago as it was suggested [26]. And now we can observe its high prevalence over other mutations in different Russian regions, from Kaliningrad in the west to Yuzhno-Sakhalinsk in the east. This mutation has also a high frequency in different European countries with a rapid decrease in frequency from east to west and in many countries in North and South America, mainly in people with European ancestry [34]. Other founder mutations (c.4035delA, c.181T>G, c.1961delA, c.68_69delAG, c.3756_3759delGTCT, and c.3700_3704delGTAAA) also had a wide distribution in Russia without any region prevalence that is similar to results of previous studies in Poland, Ukraine, Latvia, Czech Republic, and Lithuania [29,35–37]. Three of the most frequent mutations are the same as in Israel: c.68_69delAG, c.5266dupC, c.181T>G [30]. Similar results were obtained in a recent worldwide study, where 160 mutation carriers from Russia participated [29]. The data showed that c.5266dupC is the most abundant mutation followed by c.68_69delAG and c.5946delT, and the worldwide mutation frequency was highly dependent on geographic region. Some mutations identified here were observed in other countries: c.1687C>T (Austria, 14 participants in this study), c.4689C>G (Germany, 13 participants) c.4327C>T (Canada, 3 participants), c.5503C>T (Australia, 4 participants), and the common ancestral origin can be suggested. Interestingly, 54.2% of ovarian cancer patients who participated in this study contained the mutation in the BCCR versus 30.2% with a mutation in the OCCR that indicates that these regions have more speculative than clinical significance [19]. To our knowledge, we identified 89 novel pathogenic and likely pathogenic mutations in *BRCA1* and *BRCA2* genes in 113 participants, which was 30% of the total number of unique pathogenic variants detected and 6% of all participants with mutations. In recent studies, the percentage of patients with novel mutation identified was 7–19% [9,38,39]. This means that the most frequent mutations are already known in Russia but the whole set of all possible pathogenic mutations in *BRCA1/2* genes cannot be likely ever observed in real patients. However, we could reveal a new highly recurrent variants including *BRCA2* c.5286T>G that was previously identified in ovarian cancer patient and thought to be rare [40]. The haplotype analysis of these mutation carriers showed that c.5286T>G appeared to have arisen twice as late as 5382insC mutation, but more mutation carriers are necessary to unravel the time and place of its origin.

Such a high number of novel mutations observed in new studies suggests that positive selection in *BRCA1* and *BRCA2* genes may still be operating on these genes [41]. In addition to clarifying the founder pathogenic variant frequencies, we have identified several new mutations which occurrence exceeded the *BRCA2* c.5946delT mutation frequency (c.5152+1G>T, c.1687C>T, c.4689C>G, c.1510delC, and c.2285_2286delGA in the *BRCA1* gene; and c.5286T>G, c.2808_2811delACAA, c.658_659delGT, c.7879A>T, and c.3847_3848delGT in the *BRCA2* gene). Therefore, it could be useful to include them in the prescreening qPCR panel test that is widely used in Russia due to the high occurrence of several founder mutations. Then the total frequency of all mutations tested with qPCR would be 73%. In addition, we identified a novel previously unknown mutation *BRCA1* c.2285_2286delGA that was detected in eight participants. At the same time, such a high frequency of unique mutations indicates the importance of sequencing the whole coding sequences of the *BRCA1/2* genes.

One of the limitations of this study was the absence of CNV data for the Russian population because in some countries, large rearrangements (mainly equal to copy number variations, CNVs) are known to be recurrent in *BRCA1/2* genes, e.g. in Mexica (*BRCA1* ex9-12del) [42], and this limitation should be eliminated in the future.

In conclusion, this study showed the real pathogenic and likely pathogenic germline mutation occurrence in *BRCA1* and *BRCA2* genes in the Russian population; revealed new founder mutations, suggested the time of *BRCA2* c.5286T>G origin; and discovered 89 novel clinically significant variants.

## Data Availability

All data generated from the study are available as supplementary tables

## Acknowledgments

The study was supported partially under Russian State-funded budget project 0245-2021-0006 “Fundamentals of Health Preservation” and within the state assignment of the Ministry of Science and Higher Education of the Russian Federation for RCMG.

## Notes

### Competing Interest Statement

The authors have declared no competing interest.

### Funding Statement

The study was supported partially under Russian state-funded budjet project 0245-2021-0006 "Fundamentals of Health Preservation" and within the state assignment of the Ministry of Science and Higher Education of the Russian Federation for RCMG.

### Author Declarations

The study was approved by the local medical ethics committee of the Institute of Chemical Biology and Fundamental Medicine of the Siberian Branch of the Russian Academy of Sciences

## References

1. Amin N, Chaabouni N, George A. Genetic testing for epithelial ovarian cancer. Best Pract Res Clin Obstet Gynaecol. Baillière Tindall; 2020;65: 125–138. doi:10.1016/J.BPOBGYN.2020.01.005

2. George A, Kaye S, Banerjee S. Delivering widespread BRCA testing and PARP inhibition to patients with ovarian cancer [Internet]. Nature Reviews Clinical Oncology. Nature Publishing Group; 2017. pp. 284–296. doi:10.1038/nrclinonc.2016.191

3. Boussios S, Abson C, Moschetta M, Rassy E, Karathanasi A, Bhat T, et al. Poly (ADP-Ribose) Polymerase Inhibitors: Talazoparib in Ovarian Cancer and Beyond. Drugs R D. Springer; 2020;20: 55–73. doi:10.1007/s40268-020-00301-8

4. Santonocito C, Rizza R, Paris I, Marchis L De, Paolillo C, Tiberi G, et al. Spectrum of Germline BRCA1 and BRCA2 Variants Identified in 2351 Ovarian and Breast Cancer Patients Referring to a Reference Cancer Hospital of Rome. Cancers (Basel). Multidisciplinary Digital Publishing Institute (MDPI); 2020;12. doi:10.3390/CANCERS12051286

5. You Y, Li L, Lu J, Wu H, Wang J, Gao J, et al. Germline and Somatic BRCA1/2 Mutations in 172 Chinese Women With Epithelial Ovarian Cancer. Front Oncol. Frontiers Media SA; 2020;10. doi:10.3389/FONC.2020.00295

6. Kim H, Cho D-Y, Choi DH, Choi S-Y, Shin I, Park W, et al. Characteristics and spectrum of BRCA1 and BRCA2 mutations in 3,922 Korean patients with breast and ovarian cancer. Breast Cancer Res Treat. Springer; 2012;134: 1315–1326. doi:10.1007/s10549-012-2159-5

7. Janavičius R, Rudaitis V, Mickys U, Elsakov P, Griškevičius L. Comprehensive BRCA1 and BRCA2 mutational profile in Lithuania. Cancer Genet. Cancer Genet; 2014;207: 195– 205. doi:10.1016/j.cancergen.2014.05.002

8. Kim YC, Zhao L, Zhang H, Huang Y, Cui J, Xiao F, et al. Prevalence and spectrum of BRCA germline variants in mainland Chinese familial breast and ovarian cancer patients. Oncotarget. Impact Journals, LLC; 2016;7: 9600–12. doi:10.18632/oncotarget.7144

9. Heramb C, Wangensteen T, Grindedal EM, Ariansen SL, Lothe S, Heimdal KR, et al. BRCA1 and BRCA2 mutation spectrum - an update on mutation distribution in a large cancer genetics clinic in Norway. Hered Cancer Clin Pract. BioMed Central; 2018;16: 3. doi:10.1186/s13053-017-0085-6

10. Wiesman C, Rose E, Grant A, Zimilover A, Klugman S, Schreiber-Agus N. Experiences from a pilot program bringing BRCA1/2 genetic screening to the US Ashkenazi Jewish population. Genet Med. Genet Med; 2017;19: 529–536. doi:10.1038/gim.2016.154

11. Sokolenko AP, Sokolova TN, Ni VI, Preobrazhenskaya E V., Iyevleva AG, Aleksakhina SN, et al. Frequency and spectrum of founder and non-founder BRCA1 and BRCA2 mutations in a large series of Russian breast cancer and ovarian cancer patients. Breast Cancer Res Treat. 2020;184. doi:10.1007/s10549-020-05827-8

12. Kechin A, Khrapov E, Boyarskikh U, Kel A, Filipenko M. BRCA-analyzer: Automatic workflow for processing NGS reads of BRCA1 and BRCA2 genes. Comput Biol Chem. 2018;77: 297–306. doi:10.1016/j.compbiolchem.2018.10.012

13. Wang K, Li M, Hakonarson H. ANNOVAR: functional annotation of genetic variants from high-throughput sequencing data. Nucleic Acids Res. Oxford University Press; 2010;38: e164. doi:10.1093/nar/gkq603

14. Robinson JT, Thorvaldsdóttir H, Winckler W, Guttman M, Lander ES, Getz G, et al. Integrative genomics viewer. Nat Biotechnol. NIH Public Access; 2011;29: 24–6. doi:10.1038/nbt.1754

15. Kechin A, Borobova V, Boyarskikh U, Khrapov E, Subbotin S, Filipenko M. NGS-PrimerPlex: High-throughput primer design for multiplex polymerase chain reactions. Pertea M, editor. PLOS Comput Biol. Public Library of Science; 2020;16: e1008468. doi:10.1371/journal.pcbi.1008468

16. 1000 Genomes Project Consortium {fname}, Abecasis GR, Auton A, Brooks LD, DePristo MA, Durbin RM, et al. An integrated map of genetic variation from 1,092 human genomes. Nature. 2012;491: 56–65. doi:10.1038/nature11632

17. Browning SR, Browning BL. Rapid and accurate haplotype phasing and missing-data inference for whole-genome association studies by use of localized haplotype clustering. Am J Hum Genet. University of Chicago Press; 2007;81: 1084–1097. doi:10.1086/521987

18. Gandolfo LC, Bahlo M, Speed TP. Dating rare mutations from small samples with dense marker data. Genetics. Genetics; 2014;197: 1315–1327. doi:10.1534/genetics.114.164616

19. Rebbeck TR, Mitra N, Wan F, Sinilnikova OM, Healey S, McGuffog L, et al. Association of type and location of BRCA1 and BRCA2 mutations with risk of breast and ovarian cancer. JAMA - J Am Med Assoc. American Medical Association; 2015;313: 1347–1361. doi:10.1001/jama.2014.5985

20. Janavičius R. Founder BRCA1/2 mutations in the Europe: implications for hereditary breast-ovarian cancer prevention and control. EPMA J. Springer; 2010;1: 397. doi:10.1007/S13167-010-0037-Y

21. Karami F, Mehdipour P. A Comprehensive Focus on Global Spectrum of BRCA1 and BRCA2 Mutations in Breast Cancer. Biomed Res Int. 2013;2013: 1–21. doi:10.1155/2013/928562

22. Iyevleva AG, Suspitsin EN, Kroeze K, Gorodnova T V., Sokolenko AP, Buslov KG, et al. Non-founder BRCA1 mutations in Russian breast cancer patients. Cancer Lett. 2010;298: 258–263. doi:10.1016/j.canlet.2010.07.013

23. Infante M, Duran M, Acedo A, Sanchez-Tapia EM, Diez-Gomez B, Barroso A, et al. The highly prevalent BRCA2 mutation c.2808_2811del (3036delACAA) is located in a mutational hotspot and has multiple origins. Carcinogenesis. Oxford Academic; 2013;34: 2505–2511. doi:10.1093/carcin/bgt272

24. Jakimovska M, Kostovska IM, Popovska-Jankovic K, Kubelka-Sabit K, Karadjozov M, Stojanovska L, et al. BRCA1 and BRCA2 germline variants in breast cancer patients from the Republic of Macedonia. Breast Cancer Res Treat 2018 1683. Springer; 2018;168: 745–753. doi:10.1007/S10549-017-4642-5

25. Kluz T, Jasiewicz A, Marczyk E, Jach R, Jakubowska A, Lubiński J, et al. Frequency of BRCA1 and BRCA2 causative founder variants in ovarian cancer patients in South-East Poland. Hered Cancer Clin Pract. BioMed Central; 2018;16: 6. doi:10.1186/s13053-018-0089-x

26. Hamel N, Feng BJ, Foretova L, Stoppa-Lyonnet D, Narod SA, Imyanitov E, et al. On the origin and diffusion of BRCA1 c.5266dupC (5382insC) in European populations. Eur J Hum Genet. Nature Publishing Group; 2011;19: 300–306. doi:10.1038/ejhg.2010.203

27. Sokolenko AP, Bogdanova N, Kluzniak W, Preobrazhenskaya E V., Kuligina ES, Iyevleva AG, et al. Double heterozygotes among breast cancer patients analyzed for BRCA1, CHEK2, ATM, NBN/NBS1, and BLM germ-line mutations. Breast Cancer Res Treat. 2014;145: 553–562. doi:10.1007/s10549-014-2971-1

28. Snigireva G, Rumyantseva V, Novikova E, Novitskaya N, Telysheva E, Khazins E, et al. Algorithm of molecular genetic investigation to identify hereditary BRCA-associated breast cancer. Alm Clin Med. 2019;47. doi:10.18786/2072-0505-2019-47-002

29. Rebbeck TR, Friebel TM, Friedman E, Hamann U, Huo D, Kwong A, et al. Mutational spectrum in a worldwide study of 29,700 families with BRCA1 or BRCA2 mutations. Hum Mutat. John Wiley and Sons Inc.; 2018;39: 593–620. doi:10.1002/humu.23406

30. Barnes-Kedar I, Bernstein-Molho R, Ginzach N, Hartmajer S, Shapira T, Magal N, et al. The yield of full BRCA1/2 genotyping in Israeli high-risk breast/ovarian cancer patients who do not carry the predominant mutations. Breast Cancer Res Treat. Springer; 2018;172: 151–157. doi:10.1007/s10549-018-4887-7

31. Machackova E, Foretova L, Lukesova M, Vasickova P, Navratilova M, Coene I, et al. Spectrum and characterisation of BRCA1 and BRCA2deleterious mutations in high-risk Czech patients with breast and/or ovarian cancer. BMC Cancer. BioMed Central; 2008;8: 140. doi:10.1186/1471-2407-8-140

32. Solano AR, Aceto GM, Delettieres D, Veschi S, Neuman MI, Alonso E, et al. BRCA1 And BRCA2 analysis of Argentinean breast/ovarian cancer patients selected for age and family history highlights a role for novel mutations of putative south-American origin. Springerplus. 2012;1: 20. doi:10.1186/2193-1801-1-20

33. Suspitsin EN, Sherina NY, Ponomariova DN, Sokolenko AP, Iyevleva AG, Gorodnova T V., et al. High frequency of BRCA1, but not CHEK2 or NBS1 (NBN), founder mutations in Russian ovarian cancer patients. Hered Cancer Clin Pract. BioMed Central; 2009;7: 1– 7. doi:10.1186/1897-4287-7-5

34. Gomes R, Soares BL, Felicio PS, Michelli R, Netto CBO, Alemar B, et al. Haplotypic characterization of BRCA1 c.5266dupC, the prevailing mutation in Brazilian hereditary breast/ovarian cancer. Genet Mol Biol. FapUNIFESP (SciELO); 2020;43. doi:10.1590//1678-4685-gmb-2019-0072

35. Janavičius R, Rudaitis V, Feng BJ, Ozolina S, Griškevičius L, Goldgar D, et al. Haplotype analysis and ancient origin of the BRCA1 c.4035delA Baltic founder mutation. Eur J Med Genet. Elsevier Masson; 2013;56: 125–130. doi:10.1016/j.ejmg.2012.12.007

36. Nguyen-Dumont T, Karpinski P, Sasiadek MM, Akopyan H, Steen JA, Theys D, et al. Genetic testing in Poland and Ukraine: Should comprehensive germline testing of BRCA1 and BRCA2 be recommended for women with breast and ovarian cancerã Genet Res (Camb). Cambridge University Press; 2020;102. doi:10.1017/S0016672320000075

37. Kowalik A, Siołek M, Kopczyński J, Krawiec K, Kalisz J, Zięba S, et al. BRCA1 founder mutations and beyond in the Polish population: A single-institution BRCA1/2 next-generation sequencing study. PLoS One. Public Library of Science; 2018;13. doi:10.1371/journal.pone.0201086

38. Rashid MU, Muhammad N, Naeemi H, Khan FA, Hassan M, Faisal S, et al. Spectrum and prevalence of BRCA1/2 germline mutations in Pakistani breast cancer patients: Results from a large comprehensive study. Hered Cancer Clin Pract. BioMed Central Ltd.; 2019;17: 1–13. doi:10.1186/s13053-019-0125-5

39. Bu H, Chen J, Li Q, Hou J, Wei Y, Yang X, et al. BRCA mutation frequency and clinical features of ovarian cancer patients: A report from a Chinese study group. J Obstet Gynaecol Res. Blackwell Publishing; 2019;45: 2267–2274. doi:10.1111/jog.14090

40. Berlev IV, Urmancheeva AF, Imyanitov EN, Gorodnova TV, Kondratiev SV, Guseinov KD. The Clinical Course of Ovarian Cancer in a Patient with the Rare c.5286T>G (p.Y1762X) Mutation in the BRCA2 Gene. DoctorRu. NP Rusmedical Group; 2018;154: 43–46. doi:10.31550/1727-2378-2018-154-10-43-46

41. Lou DI, McBee RM, L. UQ, Stone AC, Wilkerson GK, Demogines AM, et al. Rapid evolution of BRCA1 and BRCA2 in humans and other primates. BMC Evol Biol. BioMed Central Ltd.; 2014;14: 1–13. doi:10.1186/1471-2148-14-155

42. Gallardo-Rincón D, Álvarez-Gómez RM, Montes-Servín E, Toledo-Leyva A, Montes-Servín E, Michel-Tello D, et al. Clinical Evaluation of BRCA1/2 Mutation in Mexican Ovarian Cancer Patients. Transl Oncol. Neoplasia Press, Inc.; 2020;13: 212–220. doi:10.1016/j.tranon.2019.11.003

